# Comparative Genomic Analysis of Non-typeable Haemophilus Influenzae in Children with Acute and Chronic Pneumonia versus Healthy Controls

**DOI:** 10.1101/2024.04.14.24305778

**Authors:** Deming Zhang, Wenjian Wang, Chunli Song, Tingting Huang, Hongyu Chen, Zihao Liu, Yiwen Zhou, Heping Wang

**Author notes:** **Corresponding author:** Heping Wang,.

## Abstract

Non-typeable Hemophilus Influenzae (NTHi) is a common pathogen that can cause a range of respiratory infections, including children pneumonia. However, NTHi can also be found in the upper respiratory tracts of healthy individuals and may not cause any symptoms. The transition of NTHi from a commensal to a pathogenic state is still not well understood. In this study, we aimed to investigate the genomic differences between NTHi isolated from healthy children and those with acute or chronic community-acquired pneumonia (CAP) to better understand the mechanisms underlying this transition. Genomic differences between NTHi isolated from the nasopharynx swabs of healthy children and the bronchoalveolar lavage fluids of children with acute or chronic community-acquired pneumonia (CAP) were analyzed and compared. The study used bGWAS (Bacterial Genome-Wide Association Study) analysis to identify phenotype convergence genes among the three groups and conducted gene enrichment, antibiotic resistance, and virulence factor analyses. Findings showed heterogeneity in the NTHi genomes among the three groups, and various phenotype transition genes that represent the evolution from a healthy to an acute or chronic clinical phenotype were identified. Multiple pathways were found to be involved in the pathogenicity and chronic adaptation of NTHi, including metabolism, synthetic, mismatch repair, glycolysis, and gluconeogenesis. Furthermore, the analysis indicated that antibiotic resistance genes against cephalosporin were commonly present in NTHi isolated from acute and chronic pneumonia patients. Overall, this genomic analysis of NTHi offers promising contributions toward precise clinical diagnosis and treatment.

**Importance:** Understanding the transition of Non-typeable Hemophilus Influenzae (NTHi) from a harmless commensal organism to a dangerous pathogen responsible for respiratory infections such as pneumonia in children is crucial for developing more effective diagnostic and treatment strategies. The importance of this study lies in its comprehensive examination of the genomic differences between NTHi strains found in healthy individuals and those causing acute or chronic community-acquired pneumonia (CAP). By employing advanced techniques like bGWAS (Bacterial Genome-Wide Association Study), the research sheds light on the complex genetic underpinnings that facilitate NTHi’s shift towards pathogenicity. Identifying specific genes associated with phenotype transitions, antibiotic resistance, and virulence factors not only deepens our understanding of NTHi’s biology but also paves the way for targeted therapies that could mitigate the impact of this pathogen on public health. Furthermore, the discovery of multiple pathways involved in NTHi’s adaptation to chronic infection states highlights the multifaceted nature of bacterial pathogenesis and underscores the necessity of a nuanced approach to combating these infections. This study’s findings are particularly significant given the growing concern over antibiotic resistance, as evidenced by the prevalence of cephalosporin-resistant genes in strains isolated from pneumonia patients. Thus, this research contributes importantly to the ongoing efforts to refine our approach to diagnosing and treating respiratory infections caused by NTHi, with potential implications for reducing the burden of these diseases on affected populations worldwide.

## Introduction

Childhood severe pneumonia is a serious respiratory disease with a high incidence and mortality rate among children in developing countries. To date, pneumonia remains one of the leading causes of death among all children who die before the age of five as per etiologic investigations[1, 2]. The most common pathogens causing childhood pneumonia include *Streptococcus* pneumoniae, *Haemophilus Influenza* type b (Hib), respiratory syncytial virus (RSV), and *Mycoplasma pneumoniae*[3, 4]. In the past, invasive diseases caused by *H. influenzae* were often linked to strains with type b polysaccharide capsules. However, since the introduction of the Hib conjugate vaccine, there has been a significant decrease in the frequency of illnesses caused by this particular serotype[5, 6]. Worldwide, there has been a significant increase in invasive infections caused by non-vaccine-preventable *H. influenzae* strains. This increase is mainly attributed to NTHi strains[7, 8].

NTHi is an opportunistic pathogen of the upper respiratory tract in healthy children, which can infect the lower respiratory tract and induce chronic pulmonary diseases. Studies have shown that NTHi is the most commonly isolated bacterial pathogen in otitis media and sinusitis in children and one of the major drivers of acute exacerbation of chronic obstructive pulmonary disease (COPD) in adults[9, 10]. NTHi colonization/infection is also common in young children with cystic fibrosis[11]. Currently, the molecular basis for the transformation of NTHi from a commensal organism to a pathogen is not well understood. Some studies suggest that the phase variation of NTHi is one of the mechanisms underlying its virulence changes, which include homologous recombination or changes in simple sequence repeat lengths between allele variants, phase variation mediated by slip-strand mispairing, transcriptional termination introduced by frameshift mutations, and so on[12–14]. Some studies have shown that the phase variation of specific lipooligosaccharide (LOS) biosynthesis genes plays a critical role in the transition from colonizing the human nasopharynx to invading the middle ear cavity during the course of otitis media[15]. Overall, there are relatively few genomic studies on the differences between NTHi from a commensal organism to a pathogen, partly because it is difficult to obtain pathogenic samples.

The present study aims to explore the genomic differences among (NTHi) strains isolated from the nasopharynx swabs of healthy children, bronchoalveolar lavage fluids (BALF) of children with acute or chronic pneumonia. The objective is to uncover the genomic characteristics that enable NTHi to adapt to pulmonary infections and the underlying mechanisms through which NTHi causes acute and chronic pneumonia in children. This investigation is critical for enhancing the diagnosis, treatment, and prevention of NTHi-related lower respiratory tract diseases.

## Result

### 1. Heterogeneity of NTHi genome across different clinical phenotypes

In this study, 69 samples were sequenced to explore the genomic traits of NTHi with different clinical phenotypes. The samples included 23 cases of NTHi isolated from children with acute pneumonia, 27 cases from children with chronic pneumonia, and 19 NTHi samples from the nasopharynx of healthy children for comparative analysis. All pneumonia samples (a total of 50 samples) were obtained from Bronchoalveolar Lavage Fluid (BALF), while all health control samples (19 samples) were obtained from nasopharyngeal swabs. The next-generation sequencing revealed satisfactory data quality, with an average coverage depth of 1,161x (Table S1).

While studying samples from the same patient at different clinical stages would provide a more comprehensive understanding of the changes and mechanisms of NTHi during disease progression, it presents significant challenges due to the need for sample collection before the onset of illness. This is particularly challenging in the case of children. Therefore, in the absence of the ability to obtain samples from the same patient at different clinical stages, the collection of nasopharyngeal samples from healthy individuals remains crucial for comparative analysis. Gradually increasing the sample size can help mitigate differences caused by genetic background and other factors, thereby enhancing our understanding of the biological characteristics associated with the disease.

We first attempt to classify and identify these 69 samples within the current NTHi whole-genome phylogenetic tree in order to better understand the possible relationship and evolutionary history of these samples. Determining the location of the samples in the phylogenetic tree can help us to more accurately study the evolutionary history, population structure, and genetic variation of these pathogen. Through the mining of the NTHi genomes in the National Center for Biotechnology Information (NCBI) database, the reference sequence 86-028NP was selected for the investigation of the 52 NTHi genomes filtered from the database and the 69 samples sequenced in this study. The methods used for select and filtrations are described in Method part detail. Based on the whole genomic SNP analysis, an evolutionary tree of NTHi was constructed. To the best of our knowledge, this is the most comprehensive phylogenetic tree of NTHi based on whole-genome SNPs that has been established to date (69+52=121 samples), leading to the following conclusions:

Firstly, based on the evolutionary tree analysis of the NCBI database (Fig 1B), there is significant heterogeneity in the NTHi genome within the database, which can be divided into five major branches (annotated as clade A-E in the image). Different serotypes of strains are mixed together within these NTHi branches (serotypes A, B, and D are in clade A, while serotype F is in clade E). However, this may be due to the limited representative data for each serotype, as the only three serotype A strains still cluster together relatively closely and are genetically distant from other serotypes. Previous studies have demonstrated that SNP analysis based on core and accessory genome can divide NTHi into 6 distinct clades, and that the majority of genetic information is transmitted vertically within lineages [16]. Our findings demonstrate similarities to previous studies, as notable genetic heterogeneity and lineages evolution can be observed even though we have categorized the clades into 5 groups instead of 6.

**Figure 1.**
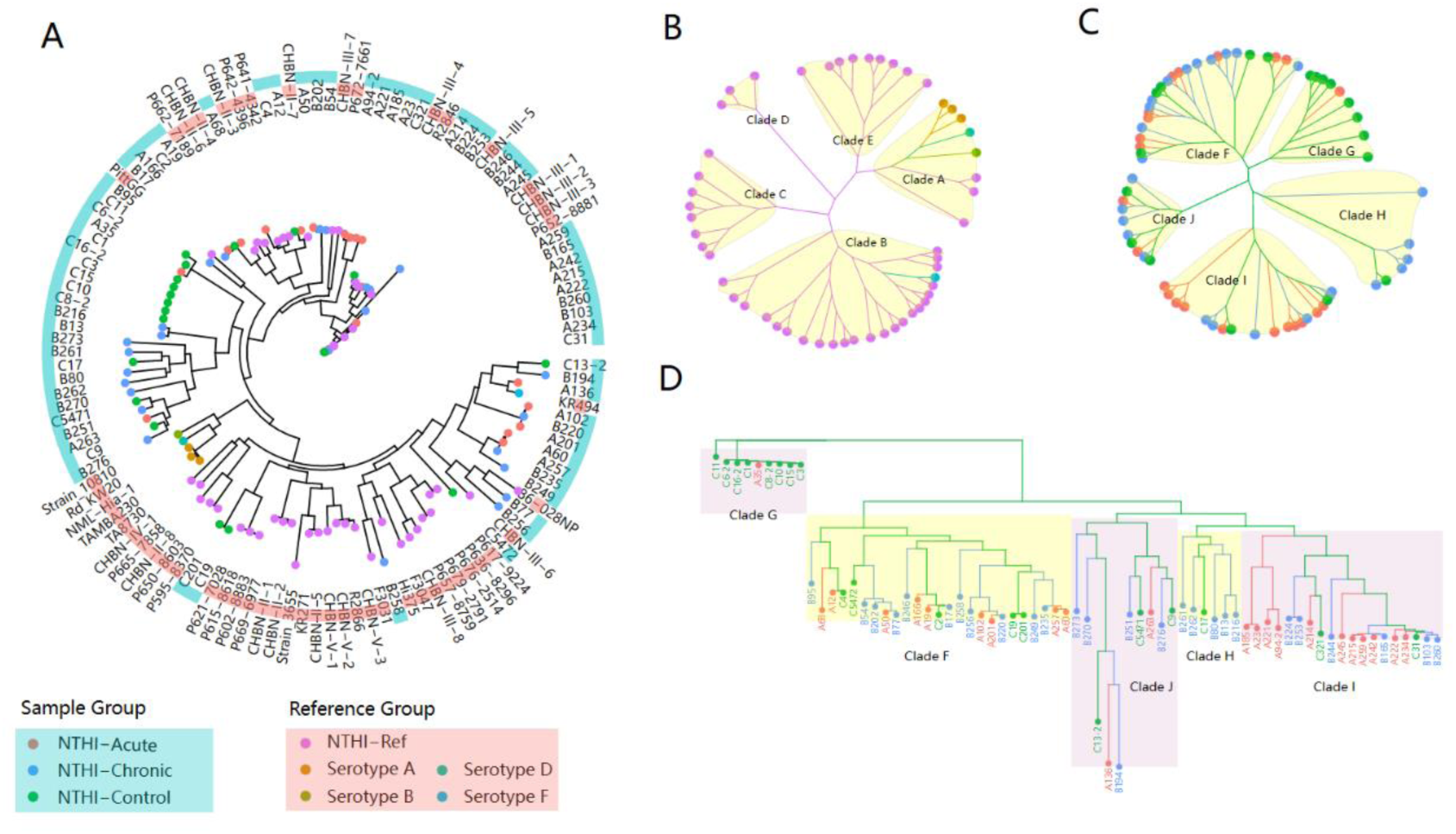
NJ(Neighbor-Joining) tree of 69 new sequence NTHi samples and 52 Hemophilus Influenza (HI) Reference Sequences in NCBI. Panel A: A combination NJ tree of samples and HI references, present as Fan layout. The outer circle of the tree is colored as red and blue, representing the reference sequences and samples, respectively. The NJ tree tip label color scheme is as follows: red for Acute cases, blue for Chronic cases, green for Healthy cases, orange for type A reference sequence, brown for type B reference sequence, teal for type D reference sequence, and light blue for type F reference sequence. B: NJ tree of 52 Influenza Hemophilus Reference Sequences, present as daylight layout. Panel C: NJ tree of 69 samples, present as daylight layout. Panel D: NJ tree of 69 samples, present as dendrogram layout, and root as Clade G. Panel B-D using the same color scheme as panel A.

As a second point, the marked heterogeneity of the NTHi genome within the database prompted us to undertake a deeper analysis of the distribution of the 69 samples in this study in relation to the 52 database reference sequences (Fig 1A, red and blue circles). These observations revealed that while a number of samples in this study clustered together (as seen in the large connected clusters of blue and red circles of Fig 1A), a cross-clustered pattern with the database sequences was still evident overall. The results suggest that the genomic differences between the samples examined in this study and those present in the NCBI database are minor, and that this study did not introduce any new clades of NTHi phylogenetic tree.

Finally, the ancestral lineage of 69 novel samples examined in this study were analyzed thoroughly, specifically regarding their evolutionary relationships. The findings of the study revealed that all the acute pneumonia (as denoted by the red color) and chronic pneumonia (as denoted by the blue color) samples under consideration, can be traced back to the same lineage as the samples of healthy children (as denoted by the green color), as depicted in Figure 1C. This phenomenon strongly suggests that NTHi pneumonia strains may have evolved from healthy strains or a common ancestor, acquiring certain traits or genetic changes that led to their pathogenic state. Despite the observation that acute pneumonia strains, chronic pneumonia strains, and healthy children’s strains do not cluster precisely in the evolutionary tree, the clustering pattern discernible in Fig 1D is evident. This clustering pattern is suggestive of the notion that genomic variations whose presence account for the clinical attributes of acute and chronic pneumonia may be complex in nature, yet the patterns of these genomic variations may still be discernible in evolutionary analysis.

### 2. Certain genes in the genomes of acute and chronic NTHi pneumonia possess distinctive phenotypic effects

Based on the above analysis, we attempted to use a method based on evolutionary tree results to analyze the correlation of whole-genome bacteria and differences in the genome of NTHi pneumonia between acute, chronic, and healthy groups. The bgwas analysis of whole-genome differences was performed using the Hogwash software [17], which captures the correlation between genome variation and phenotype variation based on the convergent evolution of the bacterial whole-genome relationship method to identify genes that can significantly affect the phenotype.

We conducted bgwas analysis on the pairwise groups of acute pneumonia vs chronic pneumonia, acute pneumonia vs Control, and chronic pneumonia vs Control. The results showed that the genes potentially determining phenotype convergence were in the range of 651-726 intervals between the three groups, accounting for between 35.5% and 39.6% of the entire reference genome of 1834 genes (Table 1). This indicates that over one-third of the genes on the genome may be involved in changes in the phenotype of health, acute pneumonia, and chronic pneumonia in this study, while most genes are relatively conservative. A total of 835 genes were identified that may be related to the dynamic evolution process of NTHi phenotype changes from health, to acute pneumonia, and finally to chronic pneumonia (TableS2).

As NTHi is not a model organism for bacterial genomic research yet, many genes of its genome do not have complete gene annotation and functional research content. We classified all the differentially expressed genes we found according to whether they could be annotated and found that only about 40% of the phenotype convergence genes found had clear functional annotation (TableS2). Other genes, usually named NTHi_geneID model genes, were genes predicted by software homology, and their function remains unclear (TableS2). As our study mainly focuses on the biological function of these phenotype convergence genes, we focus our main research efforts on genes with clearly defined functions.

The venn diagram analysis of intergroup genes shows that among these annotated genes, there are 379 phenotype convergence genes in the comparison between acute pneumonia and chronic pneumonia, which may potentially represent adaptive changes in the NTHi genome from the acute phase to the chronic phase, which we name then “Chronic adaptive genes” of NTHi in this study. There are 339 phenotype convergence genes between the acute pneumonia group and the healthy group, which may represent changes in genes from colonization to pathogenic conversion of NTHi, which we name then “pathogenic transformation genes” of NTHi in this study. Finally, we have screened a total of 274 phenotype convergence “Core genes” shared among the three groups, accounting for approximately 32.8% of the total phenotype convergence genes. These core genes mean that genes that undergo changes in phenotype from health to acute or from acute to chronic, represent the differences between colonization and infection of NTHi which are theoretically more representative than single-group comparisons (Fig 2A-C).

**Figure 2.**
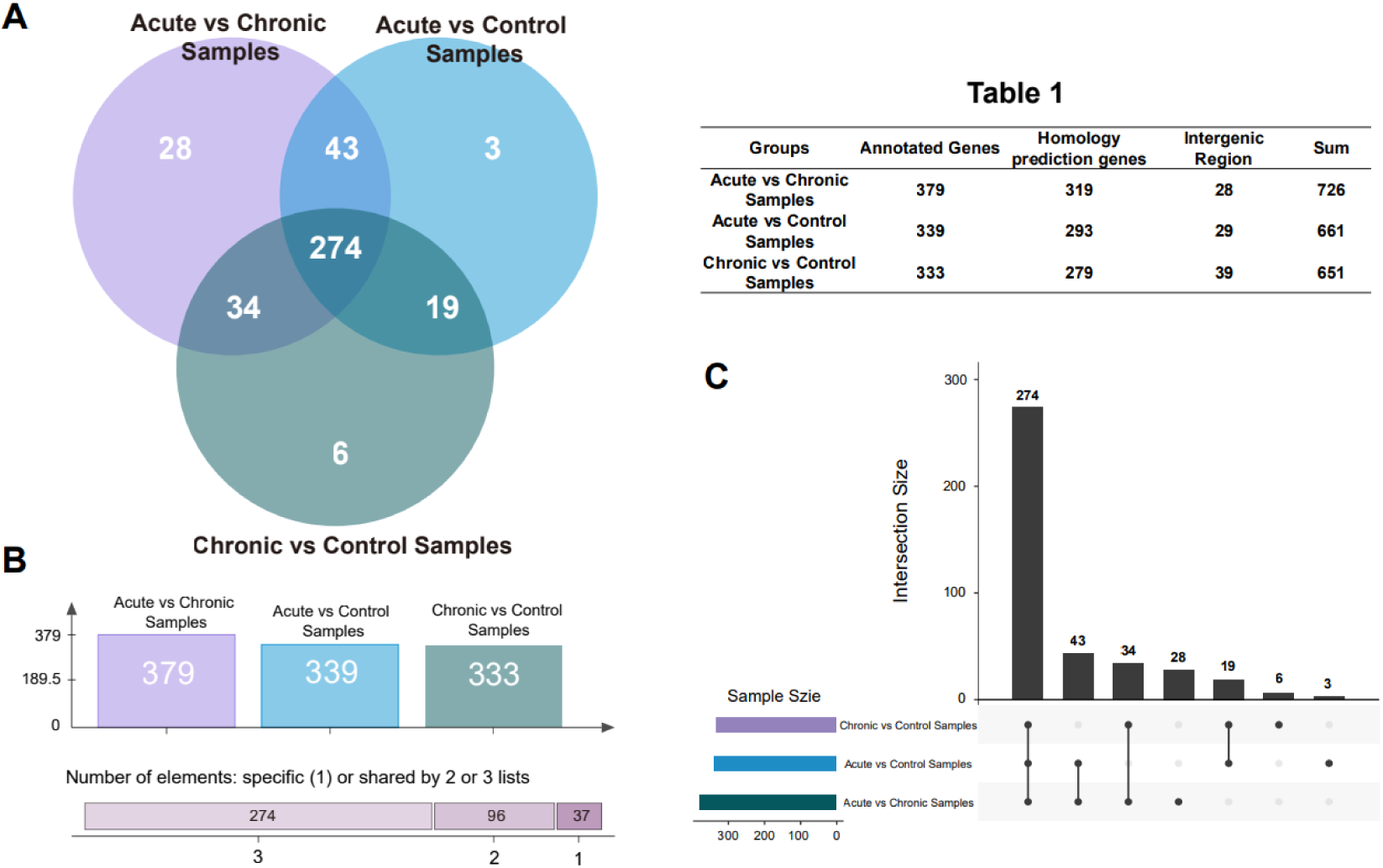
Relationship between phenotype convergence genes among different clinical groups. A. Venn diagram showing the phenotype convergence genes among acute, chronic, and healthy groups. B. Bar chart and shared relationship plot of phenotype convergence genes among the three groups. C. Upset view of phenotype convergence genes among the three groups. Table 1. Composition of annotated genes, homology-predicted genes, and intergenic regions in each clinical phenotype groups.

### 3. Multiple gene pathways involved in the Pathogenicity and Chronic adaptability of NTHi

Next, we performed enrichment analysis on the three groups of phenotype convergence genes mentioned above (Chronic adaptive genes, 379, pathogenic transformation genes, 339, Core Genes, 274). We used KOBAS-i, which is KOBAS 3.0 to perform enrichment analysis on the combination of the three selected sets of phenotype convergence genes. KOBAS gene enrichment analysis is an ORA-based (Over Represented Analysis-based) method that uses a simple and common gene enrichment method based on hypergeometric tests and Fisher’s exact tests to calculate gene enrichment pathways.

We selected a p-value threshold of <0.05 after calibration to identify significantly enriched pathways. As shown in Fig 3A-B, a total of 20 pathways were identified as significantly enriched and were classified into three categories based on their biological significance. One category is metabolism-related, shown in green in the figure, which includes nine metabolic pathways related to 1). methane metabolism, 2). c5 branch dicarboxylic acid metabolism, 3). biotin metabolism, 4). propionate metabolism, 5). starch and sucrose metabolism, 6). ketone bodies metabolism, 7). microbial metabolism under different environments, 8). carbon metabolism, and 9). metabolic pathways. Although “9). metabolic pathways” is the most significantly enriched pathway among all metabolism-related pathways based on the p-value, we should aware that its enrichment factor in the three groups of enriched pathways is only around 0.26, indicating that the pathway is statistically significant but only a small proportion of the DEGs are associated with it. On the other hand, 5). starch and sucrose metabolism and 2).c5 branch dicarboxylic acid metabolism pathways have enrichment scores of 0.67 and 0.62, respectively (TableS3), and are enriched in all three groups, indicating a high reliability that NTHi undergoes changes in the utilization of starch and sucrose and dicarboxylic acid metabolism during a clinical phenotype of colonization to chronic transformation.

**Figure 3.**
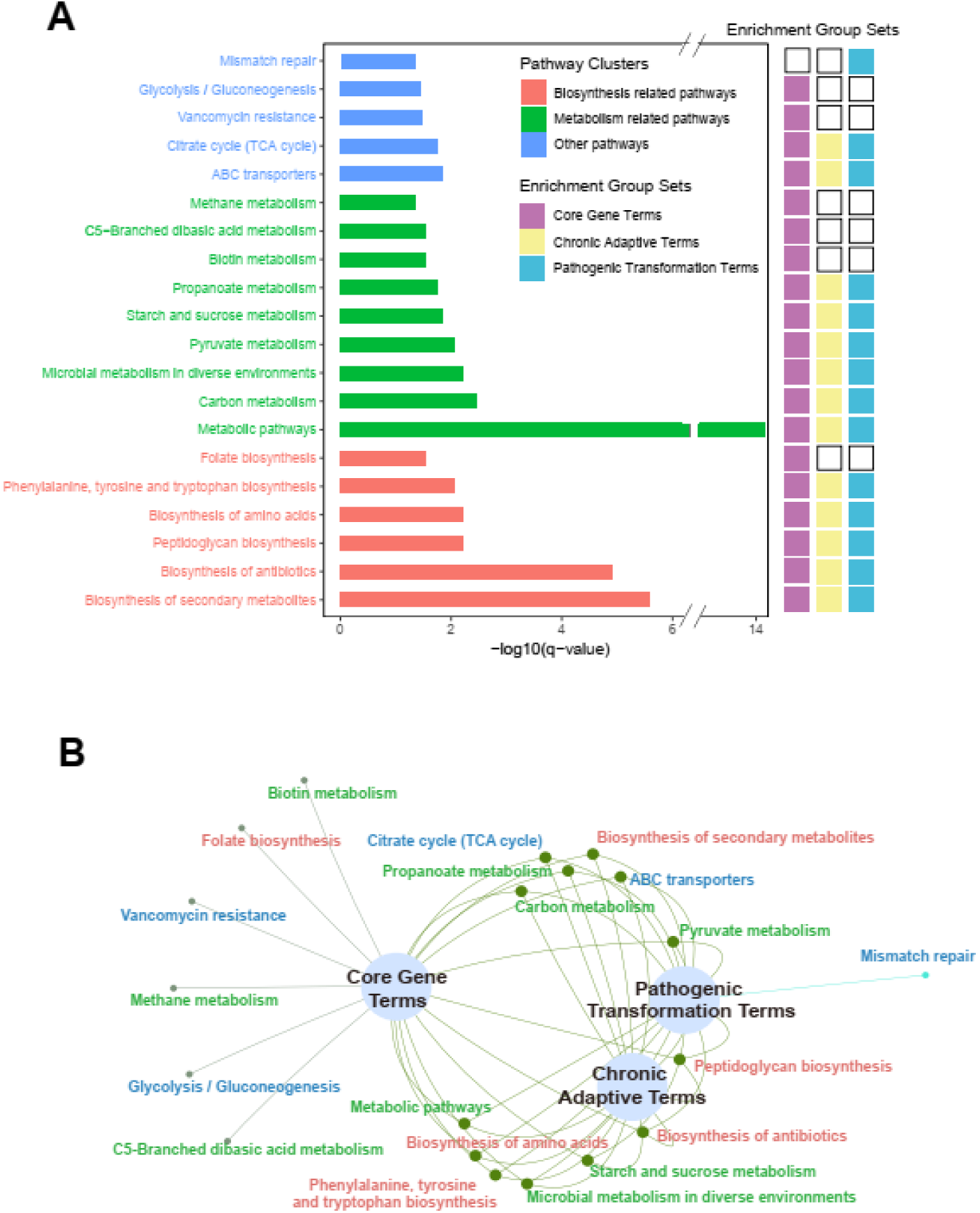
Enrichment analysis results of phenotype convergence genes in the three groups (379 Chronic adaptive genes, 339 pathogenic transformation genes, 274 Core Genes). A. Enrichment analysis results of KEGG displayed by bar plot. The results are classified into Biosynthesis-related, Metabolism-related, and Others, represented by red, green, and blue, respectively. The right panel of Figure A shows whether each term is significantly enriched in different groups. Colored fillings indicate significant enrichment, with purple representing Core genes, light yellow representing chronic adaptive genes, and light blue representing pathogenic transformation genes. B. Network diagram of the enriched terms among the phenotype convergence genes in the three groups. Common pathways are linked, and unique pathways are represented as separate branches at the end. Color scheme is the same as in A.

In addition to metabolic-related pathways, another significantly enriched pathway group is the synthetic-related pathways, which includes the 1). biosynthesis of folate, 2). phenylalanine, tyrosine, 3). tryptophan, 4). amino acids, 5). peptidoglycan, 6). antibiotics, 7). secondary metabolites, etc. Among all synthetic-related pathways, the 5). peptidoglycan biosynthesis pathway has the highest enrichment factor. Previous studies have shown that interfering with NTHi’s peptidoglycan synthesis can increase biofilm synthesis[18]; and many studies have suggested that the biofilm formed by NTHi may be an important step in the pathogenic mechanism of this bacterium [19, 20]. Our results also indicate that the peptidoglycan biosynthesis pathway, mainly composed of adaptive SNPs in the genes mraY, murA, dacB, murC, murD, murE, murF, murG, and mrcB, plays an important role in colonization and chronic transformation.

Finally, some pathways that do not belong to either the metabolic-related or synthetic-related categories were classified into the “other” category. These pathways include 1). mismatch repair, 2). glycolysis/gluconeogenesis, 3). vancomycin resistance, 4). citric acid cycle, 5). ABC transporters, etc. The 2). glycolysis/gluconeogenesis pathway is closely related to the 4). citric acid cycle, both of which are central carbon metabolic pathways that oxidize carbohydrates, fats, and proteins for energy production inside the cell. On the other hand, 5). ABC transporters play a crucial role in the pathogenic mechanism and virulence of bacteria by transferring various substrates from ions to proteins through ATP-coupled processes [21]. In addition, the 3). vancomycin resistance and 1). mismatch repair pathways were also enriched. The enrichment factor of 3). vancomycin resistance reached 0.8 in the core gene group, including four genes in this pathway, murG, alr, murF, and mraY. The 1). mismatch repair pathway was only enriched in the pathogenicity adaptive group. Overall, we found that although the number of core genes was relatively small (274 genes), they were associated with more significantly enriched pathways than those enriched in the pathogenic transformation (339 genes) and chronic adaptability (379 genes) groups. This indicates that core genes are often associated with critical pathway changes and warrants further research.

### 4. Analysis of virulence factors revealed seven specific virulence factors associated with NTHi pathogenicity

The pathogenicity of bacteria is determined by their virulence factors. Virulence factors can be encoded on mobile genetic elements (such as plasmids, genomic islands, or bacteriophages) and undergo horizontal gene transfer (propagation). Genes that control these virulence factors are present at specific sites on the chromosome, and can also be carried by genetic elements such as plasmids or bacteriophages within the bacteria. By comparing the DNA of different bacteria and identifying similar functional sequences of virulence factors, it is possible to predict the virulence of the bacteria.

It is understood that although previous studies have identified many virulence factors associated with *Haemophilus influenzae*, there has not been a comprehensive analysis and study of virulence factors of the entire genome of *Haemophilus influenzae*. In this study, we performed a whole-genome virulence factor scanning, compilation, and inter-group differential analysis of virulence factors in acute pneumonia, chronic pneumonia, and control groups based on the VFDB database. The virulence factor database, VFDB, was developed by the Chinese Academy of Medical Sciences and is widely used for virulence factor gene identification[22].

The analysis results showed that a total of seven out of eight classes of virulence factors related to *Haemophilus influenzae* were identified in 69 NTHi samples in this study (TableS4), involving aspects such as Adherence, Antiphagocytosis, Endotoxin, Protease, Immune evasion, Invasion, and Iron uptake (Fig 4B). Only the toxin-related genes, such as cdtA, cdtB, and cdtC, were not detected in any of the samples in this study. Through database analysis, we found that these not-detected virulence factors were all from the *Ducreyi Haemophilus* strain (H. ducreyi 35000HP), indicating that the different strains of *Haemophilus influenzae* have specificity. Our samples also carried some virulence factor genes of serotype D of *Haemophilus influenzae*, such as oapA, ompP5, indicating that virulence factor genes can be transmitted between different strains of *Haemophilus influenzae*. On the other hand, it is worth noting that in the chronic group, two samples were identified with a same invasion-related gene, which does not exist in the reference genome of NTHi in the database (marked as newgene in the figure). This virulence factor gene is believed to come from *Legionella*, and can enhance the invasion process of heterologous proteins.

**Figure 4.**
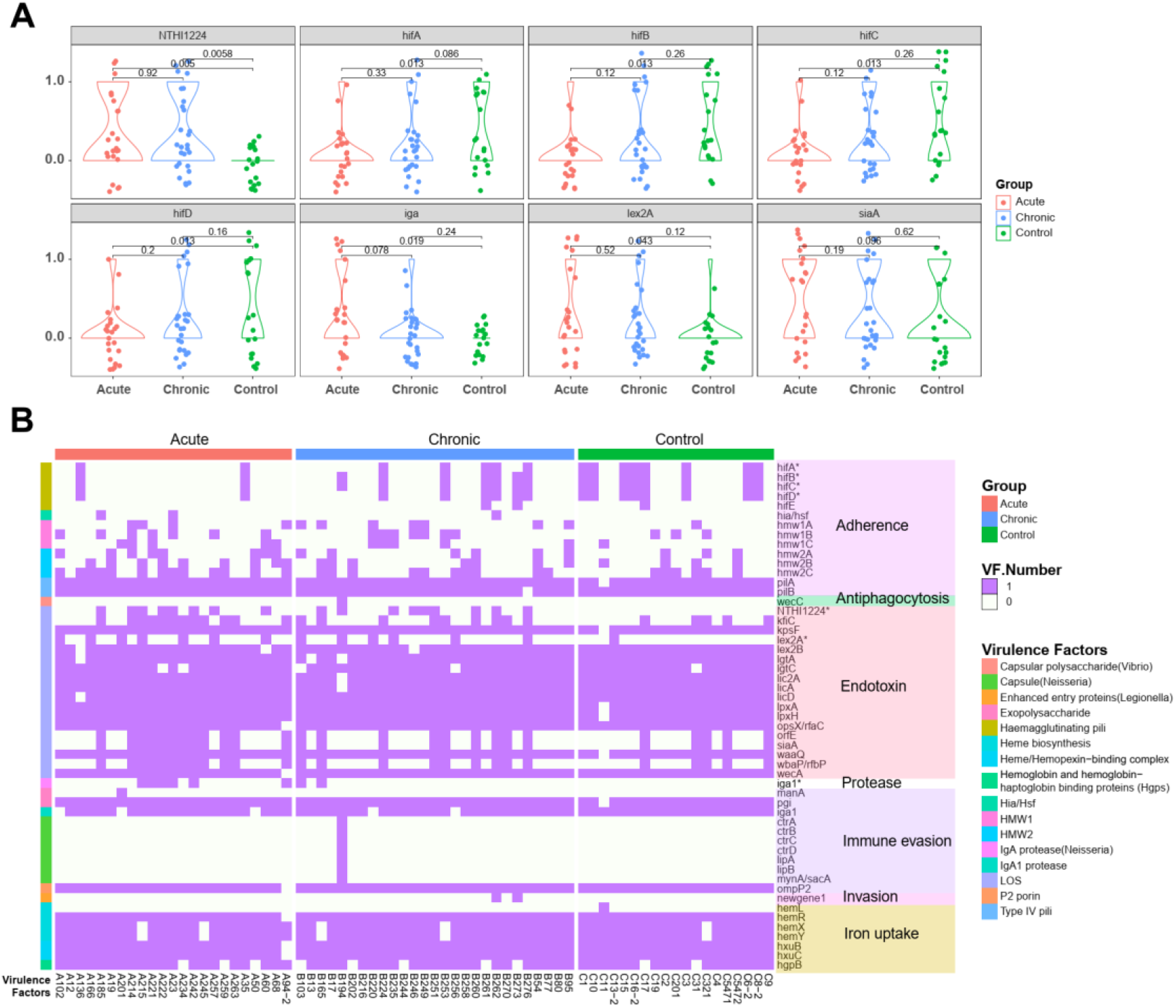
Virulence factor (VF) carriage among three clinical groups. A. Violin plots showing the difference in virulence factor carriage among three clinical groups. The *p*-value between groups was calculated using the Wilcox test. Virulence factor carriage was categorized as present or absent, with 1 indicating presence and 0 indicating absence. The points in the violin plot were jittered for better visualization. Red represents the acute group, blue represents the chronic group, and green represents the healthy group. B. Heatmap showing the virulence factor carriage among the three groups with VF classification annotations. The color fill in the heatmap represents the presence or absence of virulence factors, with purple indicating presence and white indicating absence. The first-level annotation of VF classification was directly labeled as text, and the second-level classification was represented by colored bars on the left side of the heatmap. The specific VF gene annotation was indicated in the row names of the heatmap. The colors of the three groups were the same as in Figure A.

In the end, 53 genes showing significant differences among the groups were selected for differential significance identification, which included 16 virulence factors belonging to 7 major classes. A total of 7 genes (hifA, hifB, hifC, hifD, NTHi1224, lex2A, iga) were identified as showing significant differences between any two groups. Among these, hifA, hifB, hifC, and hifD belong to the adhesion class, and their virulence factors are pili. Analysis of the inter-group differences showed that these four genes had differences between the acute and healthy groups, but the differences between the acute and chronic groups were not significant, indicating that these virulence factors may play an important role in the transition of NTHi from colonization to infection but not for NTHi chronic adaption. On the other hand, the genes NTHi1224 and lex2A belonging to endotoxins were identified, and their virulence factors were lipooligosaccharides (LOS). The LOS of NTHi is known to consist of multiple heterogenous glycan types, some of which are attributed to phase variation[23]. Previous studies have shown that LOS plays a variety of roles in serving as a colonizer and pathogen [15]. In our analysis, significant differences were found between gene NTHi1224 and the control group in both the acute and chronic groups, while differences in gene lex2A were only found between the acute and control groups. The findings suggest a partial alignment with earlier research outcomes, while simultaneously highlighting the intricate nature of biological regulatory mechanisms and the variances observed across strains.

The virulence factor of the protease class produced by the gene igA1 also showed differences between the acute and control groups, and this virulence factor is believed to be associated with the occurrence of meningitis[24]. Overall, the analysis conducted indicates that virulence factors may play a significant role in the variation of clinical phenotypes observed in NTHI. This finding may have important implications for the diagnosis, treatment, and prevention of NTHI infections, as understanding the mechanisms that underlie the pathogenicity of these bacteria is essential for developing effective interventions.

### 5. Both the acute and chronic pneumonia groups commonly carry genes for resistance to cephalosporin antibiotics

To understand the drug resistance characteristics of NTHi between different groups, we annotated all the samples involved in this study using the CARD database. The core of the CARD database is ARO (Antibiotic Resistance Ontology), which includes terms related to antibiotic resistance genes, resistance mechanisms, antibiotics, and targets. The CARD database is a widely used tool for studying resistant genes[25].

We first examined whether there were differences in ARO data identified among the three groups. The results showed that although the ARO data identified in the healthy group was lower than that in the acute and chronic pneumonia groups (with averages of 5.86 and 5.92 for acute and chronic, respectively, and 5.31 for healthy), there was no significant difference in the ARO data among the three groups overall. Furthermore, we analyzed the drug classes of the identified resistant entries in depth and found that a total of 10 drug categories were involved in the genome of this study, including elfamycin antibiotics, cephalosporins, fluoroquinolones, macrolides, monobactams, aminoglycosides, tetracyclines, nucleoside antibiotics, glycopeptide antibiotics, and sulfonamide antibiotics. Among them, more than half of the samples in this study were identified with resistance genes for five of these drugs. The most commonly identified was elfamycin antibiotic, which was detected in 57 out of 69 samples, with more than one resistance gene for elfamycin antibiotic present in 43 of these samples.

Less frequently detected resistant gene entries were mainly sulfonamide and glycopeptide antibiotics, both of which were detected only in one sample. However, drugs such as elfamycin antibiotics were detected in half of the samples in all three groups, and there was no significant difference among the groups (Fig 5A). Among the frequently detected resistance genes, the most interesting was cephalosporin antibiotics, which showed much higher detection rates in both the acute and chronic pneumonia groups than in the control group. Since the acute and chronic pneumonia samples collected in this study were all clinically treated with drugs, most of which were cephalosporin antibiotics, the genome of NTHi in the body exhibited a rapid and persistent response to drug pressure. The cephalosporin antibiotics detected in this study were divided into two categories: cephamycin cephalosporin and penam cephalosporin, with resistance gene families being OXA beta-lactamase and APH(3’’) and resistance mechanisms being antibiotic inactivation and antibiotic efflux (Table S5).

**Figure 5.**
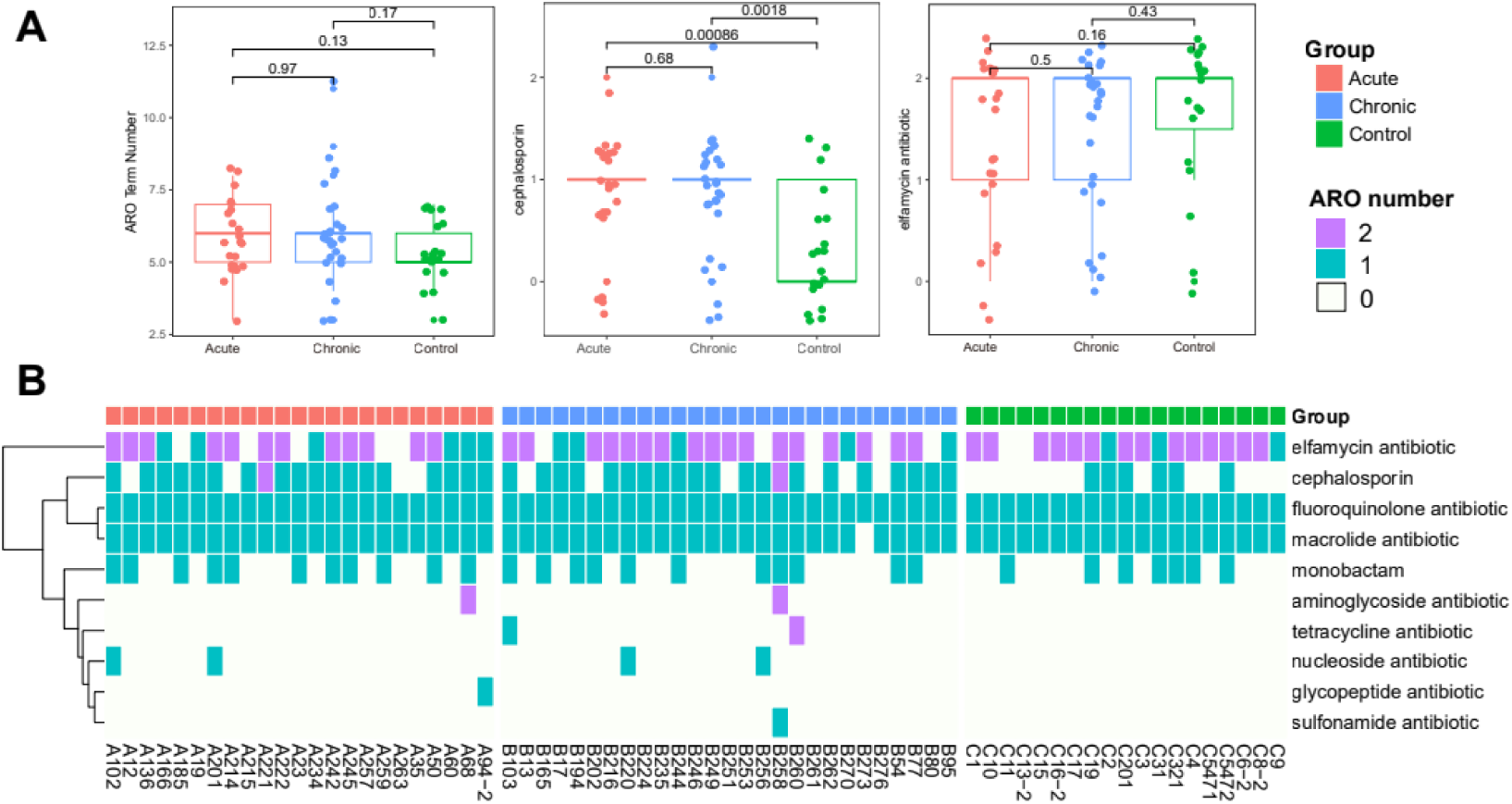
Genome-wide antibiotic resistance gene carriage in three clinical phenotypes. A. Left panel: Differential analysis of ARO numbers among three clinical phenotypes. Middle panel: Differential analysis of carriage of cephalosporin antibiotic resistance genes among three groups. Right panel: Differential analysis of carriage of elfamycin antibiotic resistance genes among three groups. *p*-values between groups were calculated using Wilcox test. B. Heatmap of antibiotic resistance gene carriage. The color in the heatmap represents the number of AROs carried for a given drug resistance type. Purple represents two copies, cyan represents one copy, and white represents no copy. Red represents the acute group, blue represents the chronic group, and green represents the healthy control group.

## Discussion

This study collected genomic sequences of NTHi from three different clinical phenotypes, including healthy children, acute pneumonia, and chronic pneumonia of children. Through comparative analysis of the genomes, the genomic characteristics of NTHi in different clinical phenotypes were explored and discussed. Firstly, by manually collating and analyzing information from the NCBI database, it was confirmed that complete NTHi genomes with serotyping are not common. The study also attempted to use whole-genome SNP analysis to differentiate NTHi genotypes and found that *influenza H. influenzae* type A, B, D, and F have potential discriminability at the whole-genome SNP level, but overall, the heterogeneity of NTHi genomes is greater. Secondly, although imperfect, there is still a certain discriminability between NTHi genomes from different clinical phenotypes. Finally, the study observed the development of NTHi from healthy to acute to chronic in the evolutionary tree, indicating that the NTHi genome can acquire convergent advantageous variations during the development of pediatric acute and chronic pneumonia.

Furthermore, we identified and distinguished convergence-associated genes through bGWAS. bGWAS analysis is a method of genome-wide association analysis based on bacterial genomes. It is a method that associates genome sequences with phenotype data to identify loci of genomic variation associated with specific phenotypes. Based on BGWAS analysis, we confirmed three representative gene enrichment analysis results. Among them, the most interesting finding was that one of our analysis results indicated that the peptidoglycan synthesis pathway may play an important role in NTHi colonization and chronic transition. Peptidoglycan is a biomolecule that plays many important biological functions in the body. Recent studies have shown that peptidoglycan synthesis also plays an important role in NTHi colonization and chronic transition. In this study, bacterial genes involved in biofilm formation were identified using a transposon mutant library and their roles in biofilm formation were verified through in vitro experiments. The experimental results of this study showed that by interfering with peptidoglycan synthesis, bacterial dissolution could be increased, leading to an increase in extracellular DNA levels and promoting biofilm formation. Interestingly, similar results could also be obtained with subinhibitory concentrations of β-lactam antibiotics, but not with other classes of antibiotics. These results suggest that the use of β-lactam antibiotics (especially NTHi strains resistant to β-lactam antibiotics) may increase biofilm formation, thereby increasing bacterial resistance to antibiotics.

Corresponding to the aforementioned findings, our analysis of NTHi resistance revealed a significant increase in the rates of cephalosporin, cephamycin, and penam resistance among the chronic and acute groups compared to the healthy group. Specifically, this resistance is due to the *flu-like haemophilus influenzae* PBP3 acquiring resistance to β-lactam antibiotics through mutations in the penicillin binding protein. The mechanism of resistance is through antibiotic target alteration. Previous epidemiological studies have shown an increasing incidence of strains with altered penicillin binding proteins, particularly PBP3 (β-lactamase-negative ampicillin-resistant and β-lactamase-positive amoxicillin-clavulanate-resistant), especially in Japan. This increase in resistance to ampicillin, amoxicillin, amoxicillin-clavulanate, and many cephalosporins has limited the effectiveness of broad-spectrum cephalosporins for meningitis and many oral cephalosporins for other diseases. Our study found that this phenomenon still exists, and our research indicates an association between this phenomenon and the formation of biofilms.

Furthermore, this study investigated the relationship between bacterial pathogenicity and virulence factors, as well as developed a method for predicting virulence based on comparative analysis of functional sequences of bacterial DNA. Building on this foundation, the study conducted a comprehensive genome-wide analysis of virulence factors in NTHi strains, revealing seven virulence factor categories related to *haemophilus* species, and identified 53 genes with differential expression between groups, including seven genes with significant differences between any two groups. These findings contribute to a deeper understanding of the virulence differences among different *haemophilus* strains and provide potential therapeutic and preventive strategies. It is worth noting that the study also identified a virulence factor gene that was not present in any known database in two samples from the chronic group, indicating the potential of this research in discovering new virulence factors.

Of course, we must acknowledge that there are certain limitations to this study, such as the incomplete recording of antibiotic use during the collection of clinical samples, as well as the time lag between sampling and drug administration. If these factors can be better controlled, we may be able to further investigate the mechanisms of antibiotic resistance and its formation in more depth. Nevertheless, overall, this study represents the first comparative genomic analysis of NTHi across multiple clinical phenotypes, with potential guiding significance for NTHi genomics and clinically related research.

## Method

### 1. Sample collection, strain identification, and identification of NTHi types

The clinical samples used in this study were obtained from the sample library of the Clinical Microbiology Laboratory at Shenzhen Children’s Hospital. The samples were collected between February 2019 and October 2021. They consisted of Haemophilus influenzae strains obtained from the nasopharynx of healthy children and bronchoalveolar lavage fluids of hospitalized children. The healthy children’s samples were taken from nasopharyngeal swabs during routine health examinations. On the other hand, the hospitalized children included both acute and chronic respiratory infection cases and underwent bronchoscopy and lavage for clinical purposes.

The children included in the study exhibited clinical symptoms of pneumonia such as coughing, chest tightness, wheezing, and fever, along with characteristic pulmonary imaging features. Those who developed these symptoms within one month were classified as having acute pneumonia, while those with symptoms persisting for more than three months were classified as having chronic pneumonia.

The lavage fluid was subjected to routine bacterial culture, with the specimens inoculated onto *Haemophilus influenzae* selective plates and Columbia blood agar plates (both purchased from bioMérieux, France) and incubated at 35°C under a 5% CO_2_ atmosphere for approximately 24 hours. After incubation, small, dewdrop-shaped, colorless, and transparent colonies were selected as suspected colonies. Gram staining was performed, with the results showing short, small, and gram-negative rods. Bacterial colonies were picked and plated onto VITEK MS-DS target plates, followed by matrix solution addition for lysis, air-drying, and on-machine analysis of the results. All isolated clinical strains were subjected to MALD-TOF (matrix-assisted laser desorption/ionization time-of-flight mass spectrometry) for mass spectrum acquisition and analyzed using the Merk VITEK MS database for microbiological identification, confirming the identity of the strains as *Haemophilus influenzae*. Total bacterial DNA was extracted from the *Haemophilus influenzae* and PCR detection of the *Haemophilus influenzae* capsule gene (bexA) was performed. B-type *Haemophilus influenzae* ATCC10211 was used as a positive control, and gel electrophoresis was used to observe the PCR products, enabling the classification of *Haemophilus influenzae*. Strains that did not show the target band were determined to be NTHi and included in this study.

This study was approved by the Ethical Committee of Shenzhen Children’s Hospital with registration number 2016013. Written informed consent for the storage and use of the BAL or NP samples for further studies was obtained from the parents or caregivers before enrollment.

### 2. Extraction, library preparation, and sequencing of the genome

The extraction of the sample genome was completed using the MGIEasy Fast Enzymatic DNA Library Prep Kit (catalog number: 940-000027-00), which is compatible with a variety of single-bacteria and meta-sample library preparations and is suitable for microbial WGS, meta-species identification, abundance determination, and assembly. Conventional cyclization reagent kits (MGI, catalog number: 1000020570) were used to cyclize the obtained PCR library to obtain single-stranded circular DNA. Conventional make DNB was performed using the make DNB reagent components in the MGI sequencing reagent kit. After the library was prepared, the MGISEQ-2000RS high-throughput sequencing reagent kit (FCL PE150) was used for genome sequencing (catalog number: 1000012555). The MGISEQ-2000RS high-throughput sequencing reagent kit uses combinatorial probe-anchor synthesis (cPAS) technology to aggregate DNA molecules and fluorescent probes on DNA nanospheres (DNBs), and high-resolution imaging systems are used to collect optical signals that are digitized to obtain high-quality and accurate sample sequence information.

Extract the genomic DNA and randomly break it into fragments, perform electrophoresis to recover DNA fragments of the required length, add adapters for cluster preparation, and finally perform sequencing. After the DNA samples are received, they are checked, and qualified samples are used to construct libraries: first, large DNA fragments are randomly fragmented into 500-800bp fragments using ultrasound methods such as Covaris or Bioruptor. The sticky ends generated by the fragmentation are repaired into blunt ends using T4 DNA Polymerase, Klenow DNA Polymerase, and T4PNK. Then, the 3’ end is extended with an “A” base so that the DNA fragment can be connected to a special adapter with a 3’ end carrying a “T” base. The target fragments are selected using electrophoresis to connect the products, and PCR technology is used to amplify the DNA fragments with adapters at both ends. Finally, qualified libraries are used for cluster preparation and sequencing.

### 3. Organizing and selecting the NTHi reference genome for the database

To determine the genomic status of NTHi in the database and select representative strains as the reference sequence for this analysis, we compiled all NTHi genomes in the existing NCBI nucleotide database and The Reference Sequence (Refseq) database. A total of 107 sequences between 1.6M and 2.4M nucleotide lengths were collected and compiled for this study. As the genome of NTHi was first sequenced and evaluated as 1.83M[26], theoretically, the sequences in our selected length range should represent complete or nearly complete NTHi sequences. We found that nine of the 107 sequences were deleted or removed from Refseq due to quality issues. Notably, the removed sequences include strains frequently mentioned in previous studies, such as NC_009566.1 (PittEE) and NC_009567.1 (PittGG) (see Appendix Table 1). Among the remaining sequences, 86 were identified as NZ - not finished WGS sequences, although many of them had titles suggesting complete genomes. One sequence was only available in GeneBank. Finally, a total of 11 sequences were identified as NC-level genomes.

We further manually curated and annotated the typing of the 107 sequences and found that most strains listed in NCBI did not adopt a unified typing standard. Previous studies have also shown that there is no unified standard for typing this bacterial sequence due to its diversity[27]. Nonetheless, most previous studies have identified the typing of NTHi through serotype, MLST typing, or capsule expression. Therefore, we finally selected 52 genomes with serotype annotations for evolutionary comparative analysis. For genome assembly with a reference genome, we used five NTHi strains at the NC (complete genome) level as reference sequences. For SNP analysis and gene annotation, to maintain consistency with previous studies, we used the 86-028NP strain as the reference sequence for analysis.

### 4. Filtering, alignment, SNP analysis, and construction of the evolutionary tree for the sequences

The downstream data from the aforementioned sequencing was first filtered using fastp software (version 0.20.1) [28] to remove adapters and low-quality sequences. Specifically, since the subsequent sequences needed to be used for assembly, we used the parameter --trim_poly_x --poly_x_min_len 10 to filter the poly sequences to reduce their impact on subsequent assembly analysis. After data filtering, we used bwa-men (Version: 0.7.17-r1188)[29] to perform sequence alignment and SNP and Indel analysis based on the selected 86-028NP reference genome. The mutect2 module in the Genome Analysis Toolkit (GATK) v4.1.1.0[30] was used for SNP and Indel analysis based on the reference genome. Even though GATK has a SNP and Indel variant calling pipeline called GATK-for-Microbes for bacterial reference genomes, we conducted tests and discovered that Mutect2 produced more reliable results. Therefore, we selected Mutect2 for our study.

After obtaining the SNPs for each sample through GATK, we used an internal script to construct a consensus sequence for each sample based on the reference genome. On the other hand, for the selected 52 complete reference genome sequences of NTHi, MUMmer4 [31] was used to call SNPs based on the reference sequence and construct the consensus sequence. Finally, we combined the consensus sequences of the samples and the reference sequences and used the software fasttree (version 2.1.10) [32] to construct the NJ evolutionary tree. After obtaining the tree file of the evolutionary tree through fastree, we used the ggtree [33] package in R software to display and beautify the evolutionary tree and used the ggtreeExtra [34], ggstar [35], and ggnewscale [36] packages to add annotations and layers to the evolutionary tree.

### 5. NTHi whole-genome association analysis and acquisition of phenotype-convergent genes

The whole-genome association analysis of NTHi was performed using the R package hogwash [17]. Based on a comparison between the Phyc and Synchronous algorithms that come with the software, the phenotype-convergent genes obtained from the Synchronous algorithm developed by the software were selected for subsequent analysis. We used the post-ancestral reconstruction grouping algorithm included in the software to perform gene- and differential-based grouping and analysis of SNPs. The SNP gene annotation was completed using SnpEff version 5.0e [37] based on the reference genome NC_007146.2, and SnpSift [38] was used to extract the relationship between SNPs and genes after gene annotation was completed. The homology-predicted functionally unknown genes in the reference genome, i.e., in 86-028NP, were labeled based on the NTHi-GeneID pattern.

In particular, some SNPs are annotated as existing in intergenic regions, so intergenic regions are also treated as independent units for testing whole-genome association. Therefore, the phenotype-convergent genes identified in this study are divided into three types for display: annotatable, homology-predicted, and intergenic regions. In this study, the phenotype refers to three clinical differentiations: acute, chronic, and healthy. After obtaining the phenotype-convergent genes, the online software EVenn [39] (http://www.ehbio.com/test/venn/#/) was used to display the differences between groups. We used two modules, Interactive Venn Diagram and Upset Plot, to show the differences in gene expression between groups from different perspectives. After uploading the raw data to EVenn to obtain the original SVG-format image, Adobe Illustrator was used to integrate the images.

### 6. Enrichment analysis of phenotype-convergent genes

The enrichment analysis of phenotype-convergent genes was performed using the Gene-list Enrichment module in the online analysis software KOBAS-i (http://kobas.cbi.pku.edu.cn/) [40], with *Haemophilus influenzae 86-028NP (nontypeable)* selected as the species. Although KOBAS-i supports four pathway databases, KEGG Pathway (K), Reactome ®, BioCyc (B), and PANTHER §, as well as GO enrichment analysis, only the KEGG Pathway database is supported for the species of NTHi. On the other hand, we attempted to use gene IDs similar to NTHi-GeneID for gene annotation in KOBAS, but none of the NTHi genes were collected in the KEGG database. Therefore, we excluded genes starting with NTHi and selected annotated genes for pathway enrichment analysis. After obtaining the enrichment results, the Gene List Enrichment Visualization module was used to obtain the raw data for pathway associations and Rich Factor for each pathway. Furthermore, these data were displayed using the pheatmap package [41] in R and the Venn Network module in EVenn [39].

### 7. Genome assembly, polishing, and annotation

Genome assembly was based on filtered data from downstream sequencing, and SPAdes (v3.13.0) [42] was used for genome reassembly. Trust-contig was set for the aforementioned five high-quality NTHi genomes starting with NC, including NC_007146.2 (86-028NP), NC_014920.1 (F3031), NC_014922.1 (F3047), NC_017451.1 (R2866), and NC_017452.1 (R2846). The kmer lengths used for assembly were the software default of 33, 55, and 77 bp, and the final result was based on the optimal N50 length. In total, around 40 scaffolds with an N50 length of approximately 11K were obtained for each sample. To further obtain a draft of the NTHi whole genome for each sample, Ragout (Reference-Assisted Genome Ordering Utility) (V2.3) [43] was used for scaffold splicing. The basic principle was to construct an A-Bruijn graph by using Cactus to align genomes and obtain colinear blocks, which were then reconstructed into a chromosome-level genome. The reference in the recipe_file was set to the single reference genome NC_007146.2 (86-028NP), and the scaffolds obtained from SPAdes were used for input. Finally, complete draft genome sequences with some N regions were obtained for all samples, and the genome sizes ranged from 1.8M to 2.2M. After completing the draft genome assembly, Prokka (rapid prokaryotic genome annotation, v1.14.6) [44] was used for annotation of the whole genome, and the kingdom level selected was Bacteria.

### 8. Toxicity and drug resistance analysis

Analysis of virulence factors was based on the VFDB (Virulence Factor Database) [22] database and was performed using the online analysis software VFanalyzer (http://www.mgc.ac.cn/cgi-bin/VFs/v5/main.cgi). The “Select genus of the genome” was set to Haemophilus, “Strain name” was set to the sample name, and “upload type” was set to “Pre-annotated DRAFT genome in GenBank format”. The input sequence was the genome draft obtained from the Ragout assembly, and the representative genome was set as NC_007146.2 (86-028NP). After online analysis was performed for all samples, the analysis results were downloaded and summarized locally. The summarized results were displayed using the R package “pheatmap” [41] for heatmap visualization, “ggboxplot” function of “ggplot2” [45] for boxplot visualization between groups, and “ggpubr” [46] for calculating the significance of differences between groups. The significance of differences between groups was determined using the Wilcox test.

Analysis of drug resistance genes was based on the CARD database (https://card.mcmaster.ca/) [25] and was performed using the main module of the Resistance Gene Identifier (RGI) software, with the search mode set to DIAMOND. After obtaining the ARO entries of resistance genes for each sample, custom scripts were used to count ARO data and drug class features for each sample. The display of statistical results was also based on the R package “pheatmap” [41] and the “ggpubr” [46] package.

## Data Availability

All data produced in the present study are available upon reasonable request to the authors

## Declarations

### Consent for publication

Not applicable

### Availability of data and materials

The data supporting the findings of this study are deposited to the CNGB Sequence Archive (CNSA) of China National GeneBank DataBase (CNGBdb) with accession number CNP0005331 (https://db.cngb.org/search/project/CNP0005331/)

## Authors’ contributions

Heping Wang, Wenjian Wang, and Hongyu Chen conceived the project and designed the experiments. Zihao Liu and Deming Zhang were responsible for the collection of mRNA samples and conducting PCR and mass spectrometry analysis. Deming Zhang, Chunli Song, and Tingting Huang carried out mRNA sequencing, GWAS (Genome-Wide Association Studies) analysis, and functional enrichment analysis. The manuscript was drafted by Deming Zhang based on feedback from all authors. Yiwen Zhou supervised the project and rigorously revised the manuscript. All authors edited and approved the final version.

### Acknowledgements

I want to thank Prof. Zuguo Zhao for guidance in my research. Our conversations inspired me to write the entire work and complete it successfully. He always made me feel confident in my skills and guided me to many important publications that were quite helpful.

